# TumorCLIP: Radiology-informed vision-language alignment for interpretable MRI-based brain tumor classification

**DOI:** 10.64898/2026.03.11.26348155

**Authors:** Yishaoying Jia, Jinfu Niu, Zhonghui Qie, Zongyu Li, Andrew Laine, Jia Guo

## Abstract

Accurate classification of brain tumors from MRI is critical for guiding clinical decision-making; however, existing deep learning models are often hindered by limited interpretability and pronounced sensitivity to hyperparameter selection, which constrain their reliability in medical settings. To address these challenges, we propose TumorCLIP, a lightweight and training-efficient vision-language framework that integrates radiology-informed text prototypes with a DenseNet-based visual encoder to support clinically meaningful semantic reasoning, fused via a Tip-Adapter mechanism. TumorCLIP does not aim to introduce a new vision-language model architecture. Instead, its contribution lies in the integration of radiology-informed text prototypes tailored to MRI interpretation, a systematic evaluation of backbone stability across diverse visual architectures, and a lightweight, training-efficient CLIP-based fusion framework designed for medical imaging applications. We first conduct a comprehensive unimodal benchmark across eight representative visual backbones (EfficientNet-B0, MobileNetV3-Large, ResNet50, DenseNet121, ViT, DeiT, Swin Transformer, and MambaOut) using a standardized optimizer and learning-rate grid search, revealing performance swings exceeding 60 percentage points depending on hyperparameter choices. DenseNet121 shows the strongest stability-accuracy trade-off within our evaluated optimizer and learning-rate grid (97.6%). Leveraging this foundation, TumorCLIP fuses image features with frozen CLIP-derived text prototypes, achieving concept-level explainability, architectural support for zero-shot and few-shot inference, and enhanced classification of minority tumor classes. On the test set, TumorCLIP attains 98.5% accuracy and reduces inter-class misclassification compared to the unimodal DenseNet121 baseline. Semantic interpretability analysis further confirms that TumorCLIP achieves a positive semantic margin rate of 96.9% across all tumor categories, compared to 42.1% for the unimodal baseline, demonstrating that its embeddings are both discriminative and radiologically meaningful. Importantly, under cross-dataset evaluation, it demonstrates improved robustness with smaller performance degradation across tumor categories, particularly for clinically heterogeneous classes such as glioma. These results suggest that the framework maintains strong predictive performance while providing an explicit radiology-informed semantic reference page. TumorCLIP therefore provides a practical, interpretable, and data-efficient alternative to conventional visual classifiers. It highlights the role of radiology-aware vision-language alignment in MRI-based brain tumor classification. All results are reported within the evaluated datasets and training protocols.

## Introduction

Magnetic resonance imaging (MRI) is indispensable for diagnosing and managing brain tumors due to its superior soft-tissue contrast and non-ionizing nature. While deep learning approaches have achieved promising results in automating tumor classification from MRI [1], clinical adoption is hampered by limited interpretability, sensitivity to hyperparameters, and the “black box” nature of vision-only architectures [2]. Such limitations undermine trust, reproducibility, and the safe application of AI in cases involving rare or visually subtle tumor phenotypes.

Recent breakthroughs in vision-language models, notably CLIP [3], have enabled powerful zero-shot learning and semantic alignment by jointly representing images and text. Although these models offer significant potential for explainable decision-making, their adoption in medical imaging and neuroimaging remains minimal. This is primarily due to a lack of large-scale paired image-text datasets and a semantic gap between natural language and the specialized descriptors used in radiology.

Existing multimodal methods also face significant barriers: paired radiology reports are scarce, radiology-specific language diverges from CLIP’s natural image training data, and end-to-end multimodal networks are often computationally intensive. Furthermore, most prior work does not provide a standardized evaluation of visual backbones, leaving the reliability of multimodal fusion architectures uncertain. To address these gaps, we conduct a rigorous, unified, and unimodal benchmark across eight popular visual backbones, including CNNs, Transformers, and state-space models, using a standardized optimizer and learning-rate grid. Our results reveal hyperparameter sensitivity exceeding 60 percentage points, with DenseNet121 [4] demonstrating the highest stability and accuracy, and thus serving as the foundation for our multimodal approach.

Building on this foundation, we propose TumorCLIP: a lightweight and interpretable vision-language framework that fuses radiology-style text prototypes with DenseNet121 via a Tip-Adapter mechanism. By utilizing a frozen CLIP text encoder to generate class-specific radiology prototypes and combining these with image-derived features through a learnable fusion weight, TumorCLIP achieves concept-level interpretability, reduced training cost, and improved detection of rare tumor classes. TumorCLIP obtains 98.5% accuracy, surpassing all unimodal baselines and providing more structured, clinically relevant decision boundaries. It also demonstrates quantifiably stronger semantic alignment with radiology-informed text prototypes than vision-only classification. Collectively, these results highlight the potential of radiology-aware vision-language alignment with prototype-based semantic analysis in MRI-based brain tumor classification.

## Results

### Backbone Benchmark

To ensure a robust foundation for subsequent multimodal fusion, we first conducted a comprehensive uni-modal benchmark to identify the most stable and high-performing visual encoder. We systematically evaluated eight representative backbones spanning convolutional networks, transformers, and state-space architectures: EfficientNet-B0 [5], MobileNetV3-Large [6], ResNet50 [7], DenseNet121 [4], ViT [8], DeiT [9], Swin Transformer [10], and MambaOut (Mamba) [11]. Each model was trained using an identical grid of optimizers (SGD, Adam) and learning rates (10*^−^*^3^, 10*^−^*^4^, 10*^−^*^5^, and 10*^−^*^6^), ensuring strict comparability across architectural paradigms.

Our results revealed pronounced sensitivity to hyperparameter selection across all models (Figure 1; [12]), with accuracy ranges exceeding 60 percentage points for several backbones. For instance, MobileNetV3-Large demonstrated performance spanning from 14.1% to 97.3%, and DenseNet121 ranged from 32.1% to 98.6%. Transformer architectures such as ViT and Swin exhibited similar volatility, although they generally surpassed CNNs at lower learning rates.

**Figure 1:**
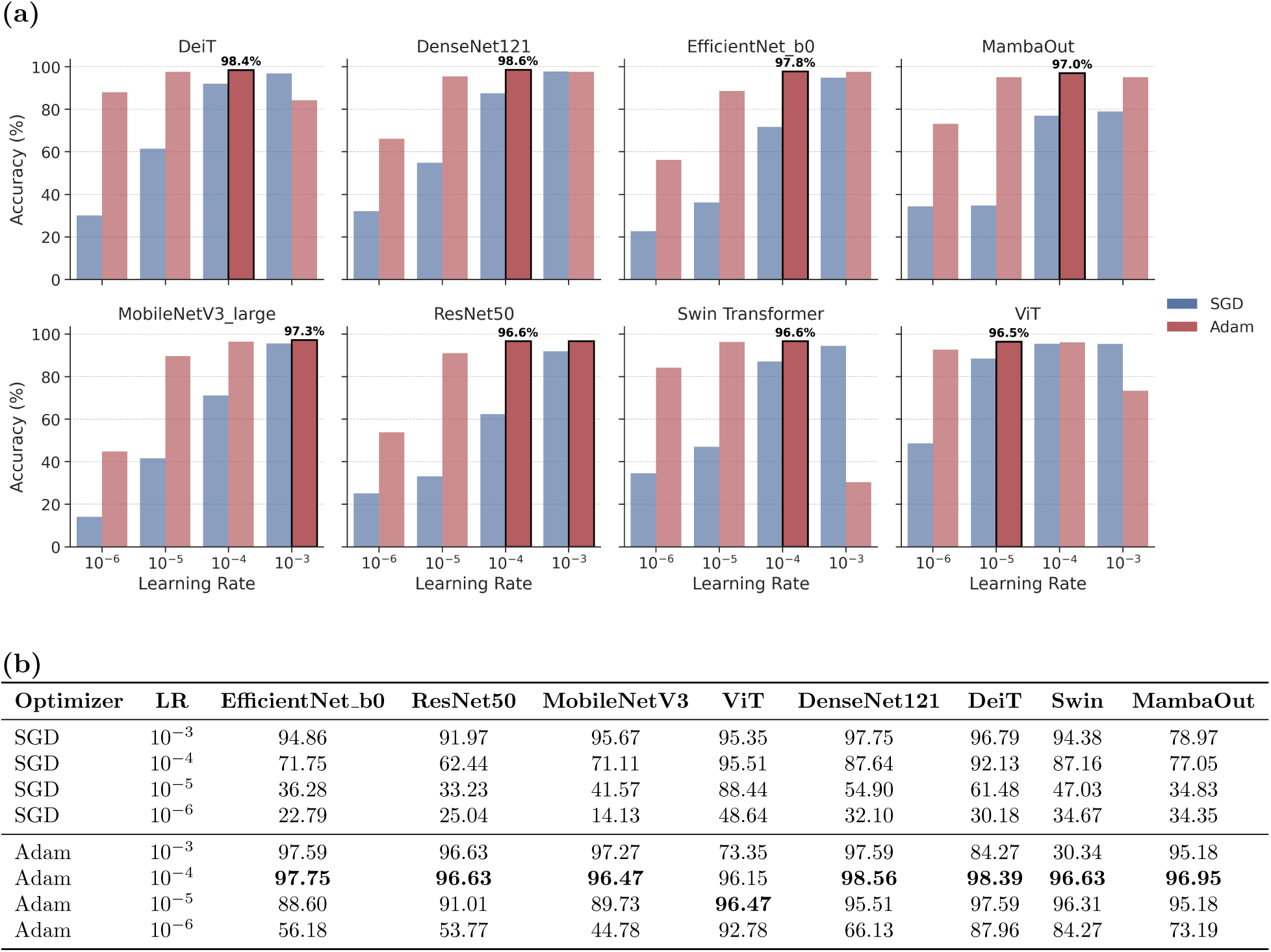
**Validation performance of single-modal models.**All visual backbones were evaluated under the same optimizer and learning-rate grid to ensure fair comparison across architectures. (a) Validation accuracy bar charts for each backbone. (b) Detailed performance table across all optimizers and learning rate configurations. The highest accuracy for each model is highlighted in bold.

Among all candidates, DenseNet121 showed the strongest reliability within the evaluated optimizer and learning-rate grid, delivering the highest validation accuracy (98.6%) and a test accuracy of 97.6% while maintaining a modest trainable parameter count (14.84M). Its robustness across diverse optimizer and learn-ing rate combinations, coupled with favorable parameter efficiency, established DenseNet121 as the optimal backbone for integration within the TumorCLIP framework.

### TumorCLIP Architecture and Multimodal Fusion

As illustrated in Figure 2, TumorCLIP consists of two parallel branches: a visual pathway and a text pathway, which are integrated through a lightweight Tip-Adapter fusion module followed by a weighted ensemble. In the visual pathway, an input MRI volume is encoded by a fine-tuned DenseNet121 backbone to produce a fixed-dimensional image embedding, together with classifier logits generated by the backbone’s classification head. In parallel, the text pathway encodes radiology-style class prompts using a frozen CLIP text encoder. For each diagnostic category, prompt embeddings are averaged to form a class-level text prototype, which serves as a semantic reference during inference. To incorporate instance-level visual evidence, TumorCLIP employs a Tip-Adapter module that leverages a precomputed cache of training-image features to supply instance-level visual evidence [13]. Given a test image embedding, the Tip-Adapter performs k-nearest-neighbor retrieval within the cache and aggregates the retrieved samples into class-specific cache scores. These cache-based scores are combined with similarity scores computed between the test embedding and the class-level text prototypes through a learnable weighting mechanism, yielding the Tip-Adapter prediction. Finally, this Tip-Adapter output is combined with the DenseNet classifier logits via a learnable weighted fusion to obtain the final inference result.

**Figure 2:**
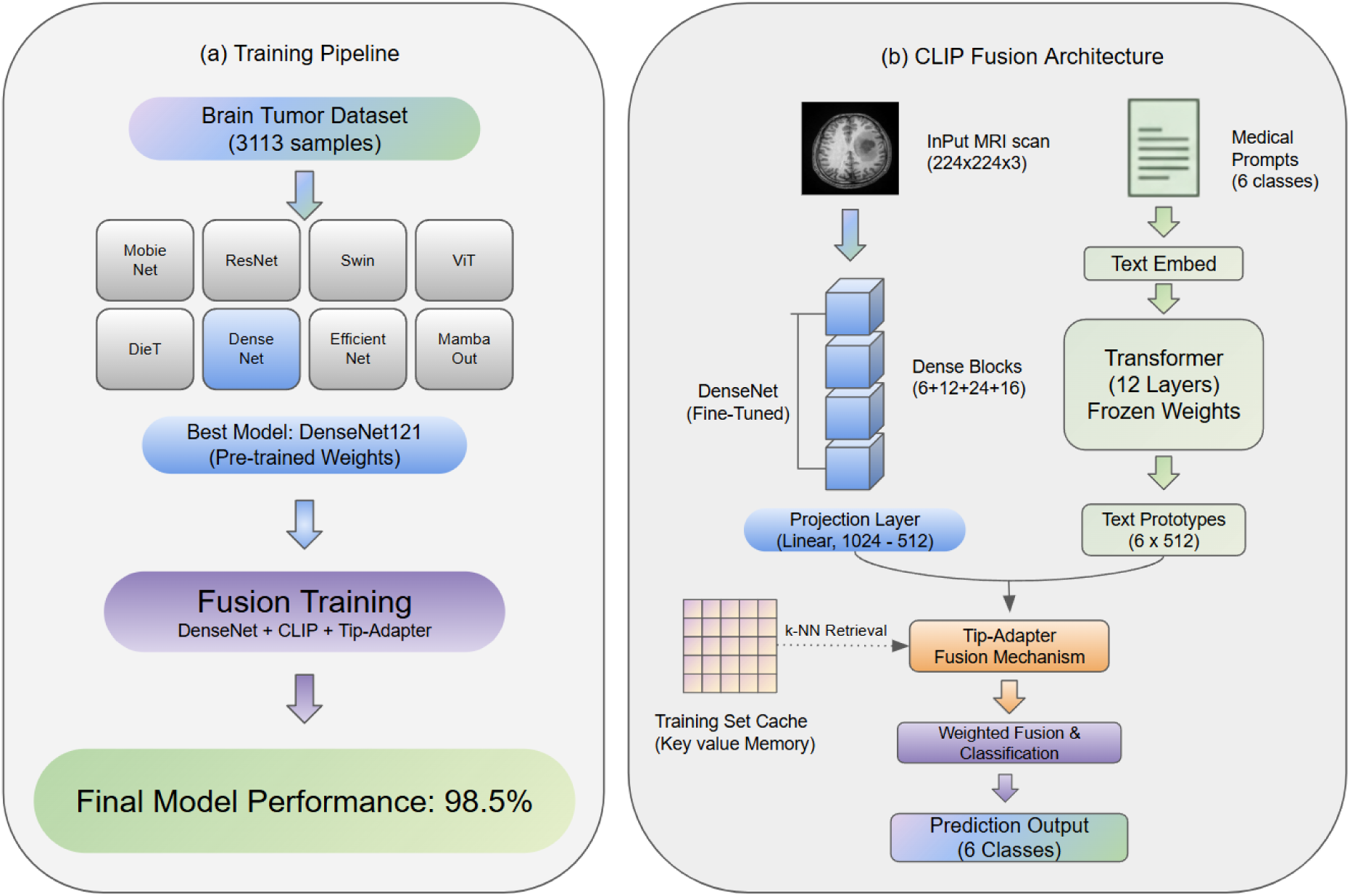
TumorCLIP training pipeline and fusion architecture. (a) Two-stage pipeline including backbone benchmarking and multimodal fusion. (b) CLIP fusion module integrating image embeddings with text prototypes using Tip-Adapter.

Collectively, these components enable TumorCLIP to integrate instance-level visual similarity with class-level radiologic semantics, resulting in a lightweight multimodal classifier that supports cache-augmented classification and prototype-based semantic interpretation, while maintaining the CLIP text encoder in a fully frozen state.

### Classification Performance of TumorCLIP

Leveraging DenseNet121 as the visual backbone, TumorCLIP achieved a test accuracy of 98.5%, outperforming the unimodal DenseNet baseline (97.6%). Notably, TumorCLIP demonstrated particular strength in underrepresented categories: as evidenced in Figure 3, recall for Neurocytoma increased by 1.86 percentage points, which is notable given the rarity and morphological diversity of this subtype. Visual analysis of the confusion matrices further indicates a reduction in inter-class misclassification. This enhancement is attributed to improved semantic alignment, in which class-level conceptual priors derived from text prototypes help distinguish subtle morphological differences.

**Figure 3:**
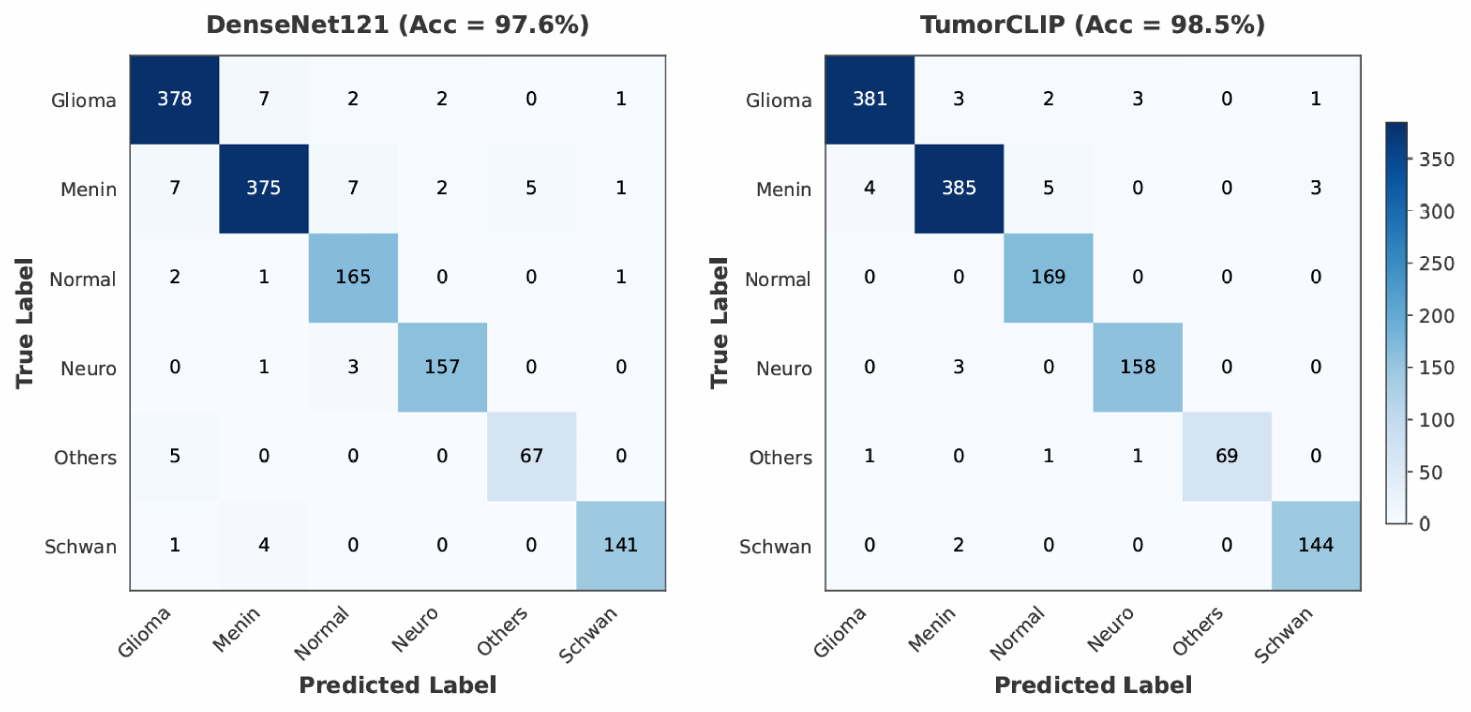
Confusion matrices of DenseNet121 and TumorCLIP. Comparison of subtype-level classification showing improved performance under TumorCLIP.

The integration of text-guided fusion sharpens decision boundaries by synergistically combining image-based features with clinically grounded radiological descriptions, thereby augmenting the model’s robustness to intraclass variability.

Crucially, TumorCLIP accomplishes these performance gains while maintaining computational efficiency. As illustrated in Figure 4, the multimodal model comprises only 14.84 million trainable parameters, which is fewer than Transformer-based baselines such as DeiT (22M), ViT (86M), and Swin Transformer (87.7M). The frozen CLIP text encoder ( 150M parameters), which remains unmodified during training, is excluded from this parameter count. Despite utilizing 5 to 6 times fewer trainable parameters than ViT or Swin, TumorCLIP achieves the highest accuracy among the evaluated models in our benchmark. This favorable parameter-accuracy trade-off positions TumorCLIP in the upper-left corner of the spectrum, underscoring its suitability for clinical applications with limited computational resources.

**Figure 4:**
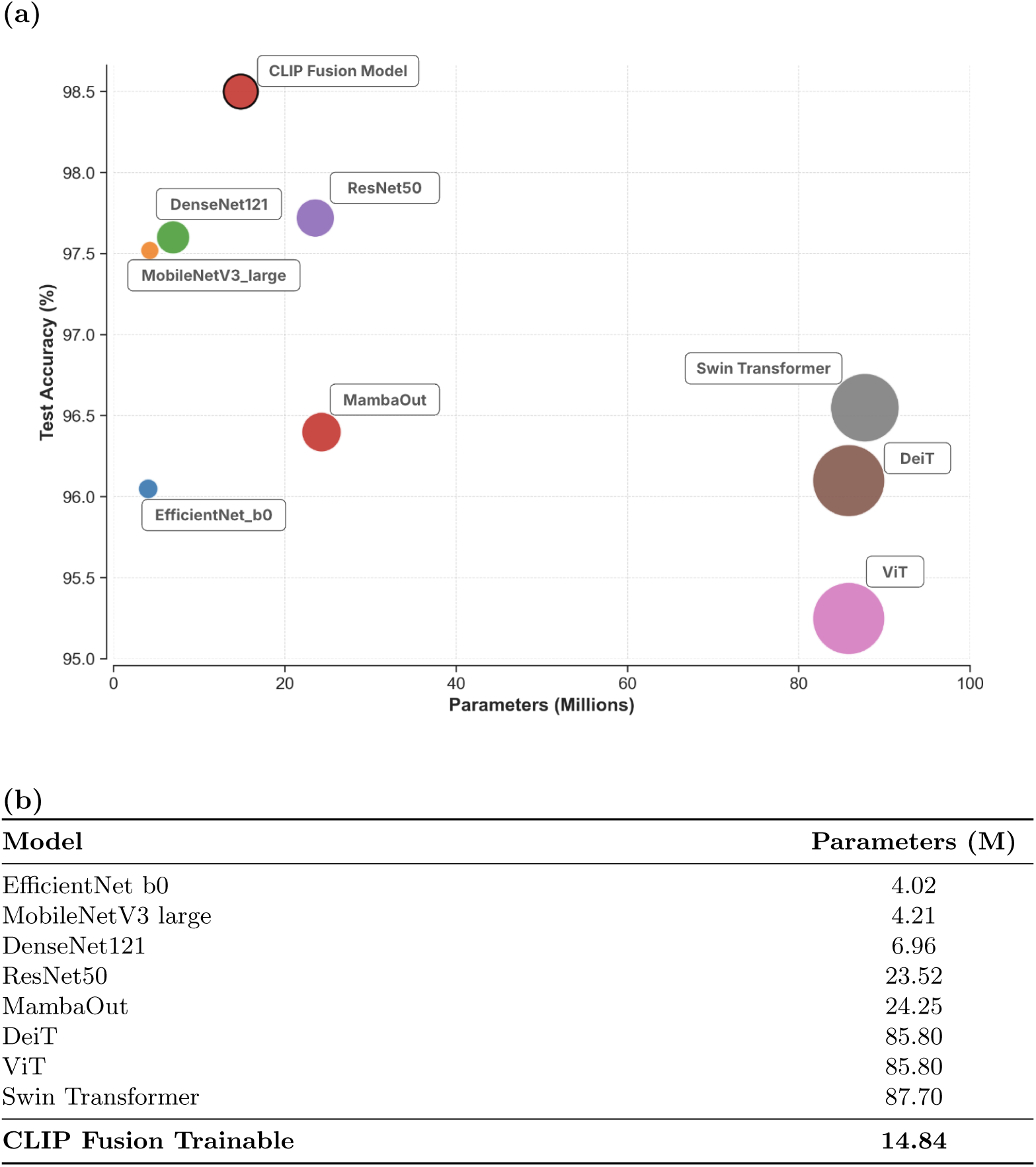
Model size and computational efficiency analysis. (a) Test accuracy versus parameter count, with bubble size proportional to the computational cost (FLOPs) of each model. (b) Detailed breakdown of trainable parameters.

### Semantic Alignment Evaluation

To assess whether learned representations align with clinically meaningful semantic concepts beyond classification accuracy, we performed a semantic interpretability analysis using text prototype similarity. For each test image, we computed the semantic margin — the cosine similarity difference between the correct class prototype and the most similar incorrect class prototype. The unimodal DenseNet121 baseline, despite achieving 96.9% classification accuracy, exhibits poor semantic consistency: a mean semantic margin near zero (-0.009) and a positive margin rate of only 42.1% (Figure 5; Table 1). This indicates that while the model’s classification head successfully separates classes, its visual embeddings are not meaningfully organized relative to radiological semantic concepts. TumorCLIP, in contrast, achieves a mean semantic margin of 0.351 and a positive margin rate of 96.9% — consistent across all tumor categories (Figure 5; Table 1).

**Figure 5:**
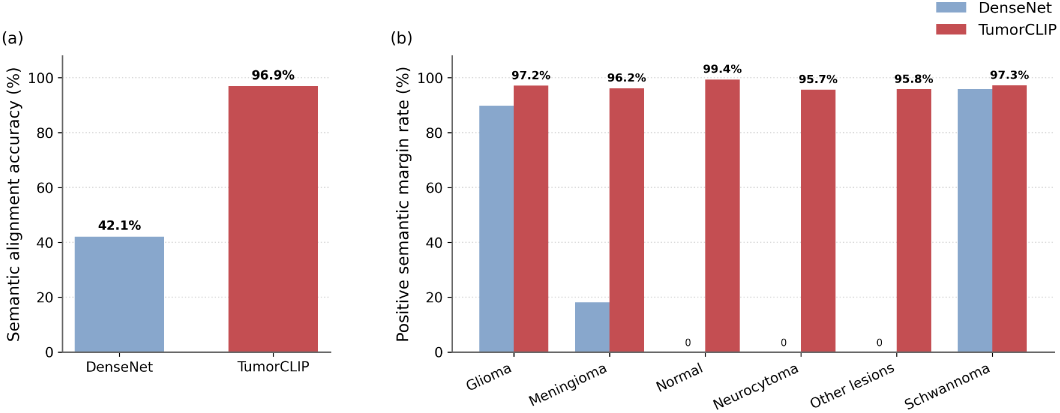
Semantic alignment performance comparison between DenseNet and TumorCLIP.

**Table 1:** Comparison of classification performance and semantic alignment between DenseNet and Tumor-CLIP.

| Model | Classification accuracy (%) | Semantic alignment accuracy (%) | Mean semantic margin | Median semantic margin |
| --- | --- | --- | --- | --- |
| DenseNet | 97.6 | 42.1 | -0.009 | -0.006 |
| TumorCLIP | 98.5 | 96.9 | 0.351 | 0.375 |

This demonstrates that the majority of image embeddings are positioned closer to their corresponding radiological text prototypes than to any alternative class, reflecting a globally coherent semantic structure in the embedding space. These results reveal a fundamental distinction between classification performance and semantic interpretability. Standard convolutional models can achieve high accuracy while maintaining internal representations that do not reflect clinically meaningful relationships. TumorCLIP explicitly aligns image features with radiology-informed textual concepts, producing embeddings that are both discriminative and semantically interpretable.

### External Dataset Generalization

To evaluate model robustness under cross-dataset variability, we further assessed DenseNet121 and TumorCLIP on an independent external MRI dataset using the trained weights without any fine-tuning. As the external dataset does not include samples corresponding to the Outros Tipos de Leso̴es category, evaluation was restricted to the five tumor classes shared with the training dataset. Performance metrics and confusion matrices were therefore computed using a filtered five-class protocol.

As shown in Figure 6, both models exhibit a performance drop when transferred from the training distribution to the external dataset. However, TumorCLIP shows improved cross-dataset performance under the evaluated distribution shift, relative to the unimodal DenseNet121 baseline. In particular, the overall accuracy degradation of TumorCLIP is smaller. This trend is further evident at the class level, where TumorCLIP maintains higher accuracy for glioma, a clinically heterogeneous category known to be particularly sensitive to variations in acquisition protocols and data sources.

**Figure 6:**
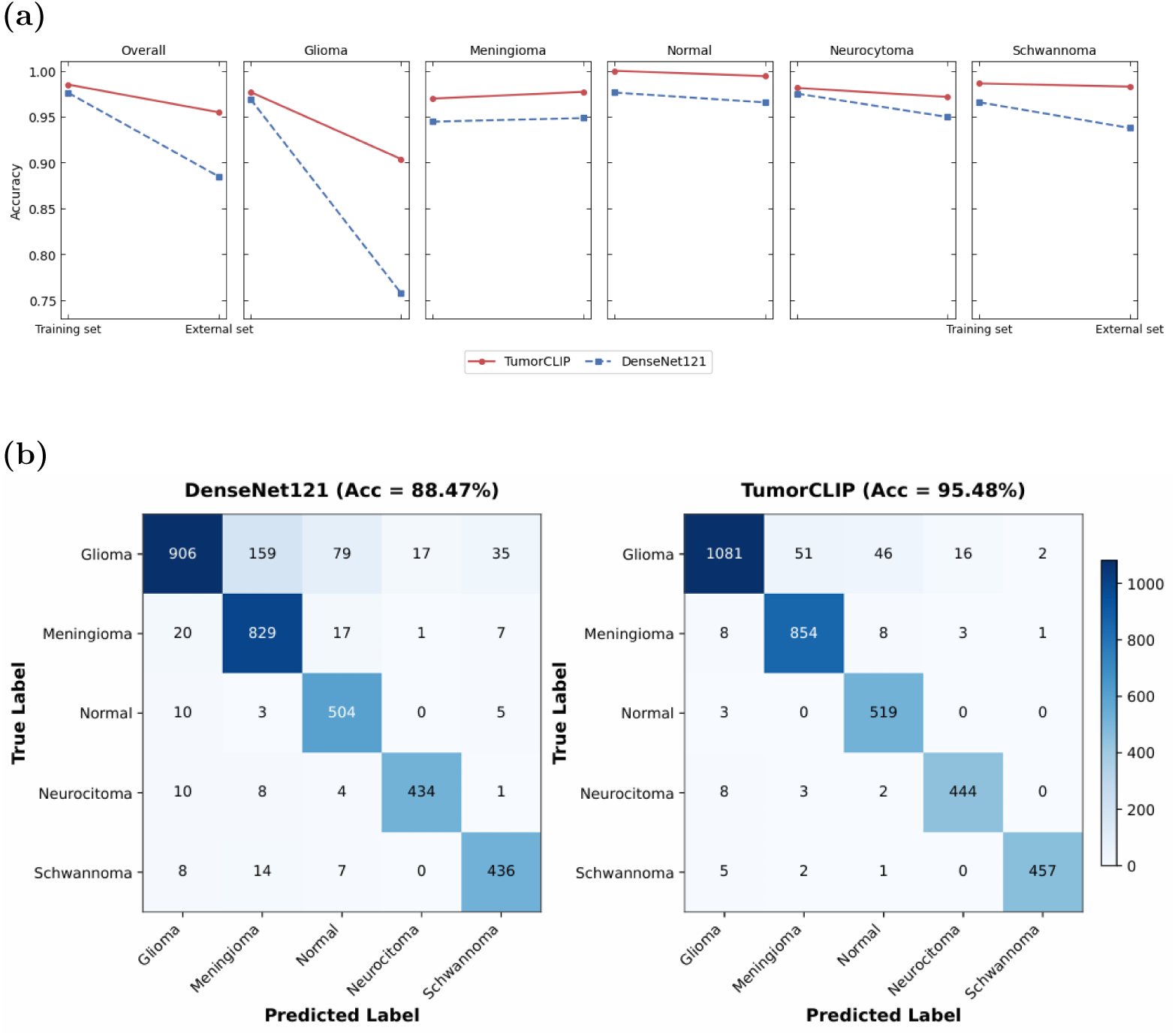
(a) Accuracy comparison between DenseNet121 and TumorCLIP on the training and external datasets across overall and tumor-specific categories. (b) Confusion matrices of DenseNet121 and Tumor-CLIP on the external dataset under a filtered five-class setting, excluding Outros Tipos de Leso̴es.

Confusion matrix analysis of the external dataset (Figure 5b) shows that TumorCLIP reduces inter-class confusion across all five evaluated categories compared to DenseNet121. Notably, misclassification between glioma and other tumor subtypes is substantially attenuated. These observations indicate better external dataset performance for the complete TumorCLIP system under the evaluated distribution shift.

### t-SNE Visualization

To qualitatively assess the impact of TumorCLIP on class separability, we visualized the model outputs using t-SNE, as shown in Figure 7. We performed t-SNE on the learned feature embeddings extracted from the last layer, enabling a direct examination of how each model structures its representation space rather than its final decision scores. This embedding-based visualization was applied consistently to both the training dataset and the independent external test dataset.

**Figure 7:**
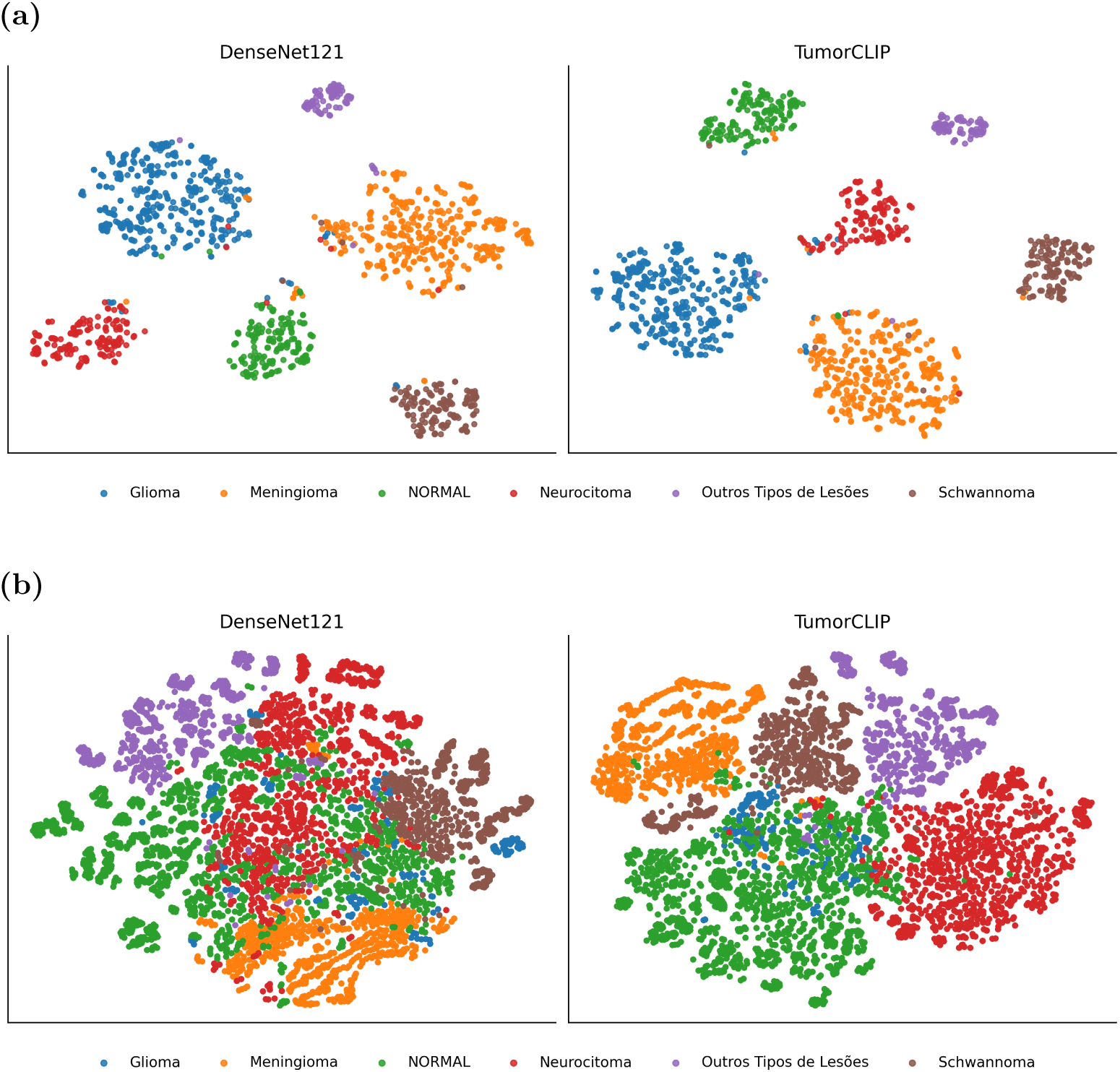
Comparative t-SNE visualization of learned feature embeddings. (a) t-SNE projections of embeddings extracted from the training dataset for DenseNet121 (left) and TumorCLIP (right). (b) Corresponding t-SNE projections of embeddings from the external test dataset. Each point represents a single sample and is colored by tumor category. TumorCLIP exhibits tighter intra-class compactness and clearer inter-class separation.

Both DenseNet121 and TumorCLIP generate six discernible clusters corresponding to the diagnostic categories in the training set. However, TumorCLIP exhibits more compact and cohesive embedding clusters, with reduced overlap at class boundaries, indicating improved class-wise organization in the learned feature space. This effect is particularly evident in the glioma cluster, where TumorCLIP forms a denser manifold with clearer separation compared to DenseNet121, consistent with its improved discrimination of visually ambiguous tumor patterns.

When evaluated on the external dataset, the contrast between the two models becomes more pronounced. DenseNet121 embeddings show increased dispersion and partial collapse between several tumor categories, reflecting reduced robustness under distribution shift. In contrast, TumorCLIP preserves a more stable geometric structure, maintaining recognizable cluster identities despite domain differences. Notably, minority tumor classes exhibit less fragmentation in the TumorCLIP embedding space, suggesting that radiology-guided semantic priors improve generalization beyond the training distribution. Overall, these embedding-level visualizations demonstrate that TumorCLIP enhances class separability not only within the training domain but also under external evaluation, yielding a more structured and resilient representation space. By injecting radiological semantics through text prototypes and cache-based fusion, TumorCLIP reshapes the embedding geometry, leading to more reliable class organization and improved robustness for rare and challenging tumor categories.

## Methods

### Model Overview

The CLIP text encoder is fully frozen throughout all experiments and is never fine-tuned. All trainable parameters are restricted to the DenseNet classifier head, the lightweight adapter, and the fusion module.

### Dataset and Preprocessing

The primary training and in-domain evaluation experiments utilized the publicly available Brain Tumor MRI Images (17 Classes) dataset from Kaggle [14]; representative examples are shown in Figure 8. To reduce extreme class imbalance and improve clinical relevance, the original 17 tumor labels were merged into six diagnostic super-classes based on lesion location and radiological appearance, including Glioma, Meningioma, Schwannoma, Neurocytoma, Normal, and Outros Tipos de Leso̴es (Supplementary Table S1). This grouping follows common radiological categorization principles and has been adopted in prior brain tumor MRI classification studies. The dataset encompasses a diverse collection of axial MRI scans, incorporating T1-weighted, contrast-enhanced T1 (T1c), and T2-weighted modalities.

**Figure 8:**
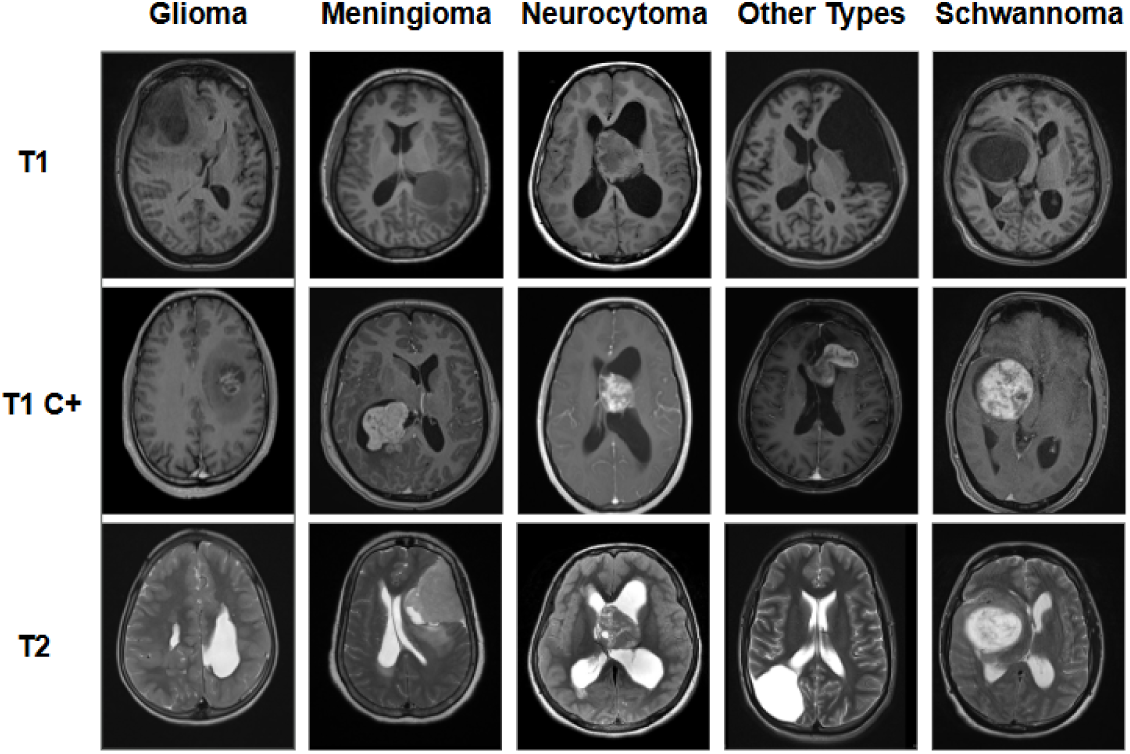
Representative axial MRI slices from five brain tumour categories. (Glioma, Meningioma, Neurocytoma, Other Types of Injuries, and Schwannoma) across T1, contrast-enhanced T1 (T1C+), and T2 modalities. Shown cases are independent examples and are not derived from the same patient across modalities.

To further evaluate model generalization, an independent external MRI dataset was used for evaluation [15]. This dataset was collected from a different source and exhibits variations in acquisition characteristics and label composition. As the external dataset does not include samples from the Outros Tipos de Leso̴es category, external performance was assessed using a filtered protocol restricted to the five tumor categories shared between the training and external datasets. During this evaluation, predictions and performance metrics were computed exclusively over these overlapping classes, with samples from the excluded category omitted from metric calculations.

Preprocessing was standardized across all models to ensure fair comparison. Each image was resized to 224 × 224 pixels, converted to a three-channel format, and normalized using ImageNet mean and standard deviation statistics. The dataset was partitioned into training, validation, and test sets via stratified sampling to maintain consistent class distributions. To enhance generalization while preserving comparability across visual backbones, we applied lightweight data augmentation limited to random horizontal flipping and minor in-plane rotations of ±10*^◦^*. No modality-specific transformations or intensity-based augmentations were employed. This unified preprocessing and augmentation pipeline was consistently used for all models throughout the study.

### Radiology Text Prompts

To infuse the multimodal branch with clinically meaningful semantics, we manually authored radiology-style text prompts for each tumor category. These prompts capture characteristic imaging phenotypes, including anatomical location, signal intensity patterns, and enhancement behavior. All prompts were encoded using a frozen CLIP text encoder, and prompt embeddings within the same category were averaged to form a robust class-level radiology text prototype.

Specifically, we constructed five radiology-style text prompts for each of Glioma, Meningioma, and Normal, and four prompts for each of Neurocytoma, Outros Tipos de Leso̴es, and Schwannoma, resulting in 27 multilingual prompts across six diagnostic categories. These radiology text prototypes serve as semantic anchors, supporting both zero-shot inference and multimodal fusion during prediction.

Representative examples of the radiology-style text prompts used to construct the class-level radiology text prototypes are shown in Table 2:

**Table 2:** Radiology text prototypes

| Class | Example radiology-style text prompt |
| --- | --- |
| Glioma | Intra-axial infiltrative lesion with heterogeneous T2 hyperintensity |
| Meningioma | Extra-axial mass with dural tail and broad dural attachment |
| Schwannoma | Enhancing mass along the cisternal segment of a cranial nerve |
| Normal | No abnormal signal intensity, mass effect, or enhancement |
| Neurocytoma | Well-defined intraventricular mass with mixed T1/T2 signal |
| Outros Tipos de Lesões | Heterogeneous lesion with irregular borders and variable enhancement |

### Unified Training Protocol and Objective Functions

To ensure a rigorously fair comparison across architectures, all eight visual backbones—EfficientNet-B0, MobileNetV3-Large, ResNet50, DenseNet121, ViT, DeiT, Swin Transformer, and MambaOut—were trained following an identical optimization protocol. Each model was evaluated using a unified hyperparameter grid that included both SGD and Adam optimizers in combination with four learning rates ranging from 1 × 10*^−^*^3^ to 1 × 10*^−^*^6^. Batch size, data preprocessing, and augmentation strategies were held constant throughout all experiments, and early stopping based on validation accuracy was employed to mitigate overfitting.

All models were trained to optimize the standard multi-class cross-entropy objective, expressed as:

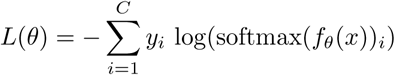

where *C* is the number of tumor classes and *f_θ_*denotes the backbone network’s forward function.

Within this standardized experimental framework, DenseNet121 consistently exhibited the highest stability and overall performance, making it the backbone of choice for the TumorCLIP architecture.

For the multimodal TumorCLIP model, we employed a composite loss function to jointly optimize the fusion logits, the DenseNet branch, and the CLIP–Tip-Adapter branch. Let **s**_fused_ denote the fused logits, **s**_dense_ the DenseNet-only logits, and **s**_clip_ the CLIP–Tip-Adapter logits. The total loss is defined as:

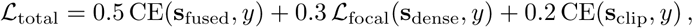

where CE represents the standard multi-class cross-entropy and L_focal_ denotes the focal loss applied to the DenseNet classifier. During multimodal training, the inclusion of a focal loss component for the DenseNet branch specifically targets hard and minority-class examples, while maintaining the cross-entropy loss used in the unimodal benchmark. This multi-task objective encourages the fusion head to remain aligned with both the unimodal DenseNet baseline and the multimodal CLIP branch, while ensuring robust learning on challenging samples. The fusion loss is assigned the highest weight to prioritize multimodal decision learning, while focal loss is applied to the DenseNet branch to improve performance on minority classes. The CLIP branch is treated as auxiliary supervision to stabilize semantic alignment during training.

### Construction of Radiology Text Prototypes

To embed clinically meaningful semantic priors within the model, we authored radiology-style textual descriptions for each diagnostic category. These prompts were designed to encapsulate characteristic MRI features commonly referenced in clinical interpretation, such as typical lesion locations (e.g., intra-axial, extra-axial, intraventricular), signal intensity patterns, enhancement characteristics, and class-specific morphological cues. Each prompt was processed through the frozen CLIP text encoder. For each class *k*, all associated text embeddings were averaged to generate a class-level text prototype:

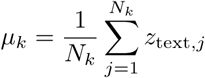

where *N_k_*is the number of prompts constructed for class *k*. Across the six diagnostic groups, each class was represented by approximately six prompts. These class prototypes serve as semantic anchors, providing reference points for aligning DenseNet-derived image features during both zero-shot inference and multimodal fusion within TumorCLIP.

### Image-feature Cache Construction

After identifying DenseNet121 as the optimal unimodal backbone, we utilized the trained network to extract feature representations from all training images, thereby constructing a non-parametric cache for use in the Tip-Adapter modules. For each training instance *x_i_*, the model generated a normalized feature embedding:

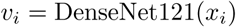

*v_i_* = DenseNet121(*x_i_*) This cache comprised two key components:

1. the collection of visual embeddings *V* = {*v_i_*}, which serve as the key space for similarity-based retrieval.
2. the corresponding one-hot label vectors *Y* = {*y_i_*}, which encode class-specific supervision signals.

During inference, this cache remains fixed and provides instance-level evidence through similarity-weighted retrieval, supporting few-shot adaptation without necessitating further training or fine-tuning of the backbone network.

### Tip-Adapter Formulation

TumorCLIP fuses semantic information from radiology-informed text prototypes with instance-level evidence from the feature cache using a lightweight adaptation strategy inspired by the Tip-Adapter framework. For each test image, the DenseNet121 backbone generates an embedding *v*, which is normalized and subsequently compared to the radiology-derived text prototypes. Prior to similarity computation, *v* is passed through a lightweight two-layer adapter MLP (512 → 128 → 512); the adapter and the image feature projection layer are jointly optimized, while the CLIP text encoder remains frozen.

Text-prototype logits are calculated via cosine similarity:

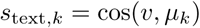

To incorporate supervision from labeled examples, the model retrieves signals from the cached training embeddings. For each cached feature *v_i_*, a similarity score is computed using cosine similarity and scaled by a temperature parameter *t*_knn_:

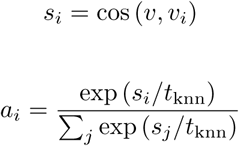

where *t*_knn_ modulates the neighborhood selectivity of the cache retrieval. Cache-based logits are then com-puted by aggregating the normalized similarity weights over the associated one-hot labels:

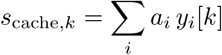

Fusion of semantic (text) and cache-based evidence is achieved through a scalar alpha: *α*:

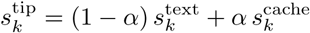

Simultaneously, the DenseNet backbone outputs conventional classifier logits for each class *k*, denoted *s*_dense*,k*_. At the model level, the Tip-Adapter logits are combined with the DenseNet logits via a learn-able fusion weight *w* ∈ (0, 1), resulting in the fused logits:

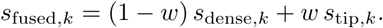

The parameter *w* is implemented as the sigmoid of an unconstrained scalar and is jointly optimized with the adapter parameters. The final class prediction is made by selecting the class corresponding to the maximal fused logit:

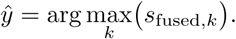

This formulation enables multiple inference modes within a unified framework. Zero-shot classification uti-lizes only the text-prototype logits *s*_text*,k*_, disabling both the cache-based logits and the DenseNet classifier head. Few-shot adaptation incorporates cache-derived logits *s*_cache*,k*_, while the complete TumorCLIP model leverages both text-prototype and cache-based evidence via a fixed mixing coefficient *α* (tuned as a hyper-parameter) and a learnable fusion weight *w*. Importantly, the CLIP text encoder remains entirely frozen throughout all stages.

The feature cache contains embeddings of all training samples extracted using the frozen image encoder. Similarity-based retrieval from the cache is controlled by a temperature parameter *t*_knn_, which corresponds to *β* = 1*/t*_knn_ in the exponential weighting formulation. In all experiments, *t*_knn_ is set to 0.07.

The lightweight adapter is trained for N = 15 epochs using the same optimizer and learning rate settings as the DenseNet classifier, while all other components, including the text encoder, remain frozen. All hyperparameters are kept fixed across experiments to ensure fair comparison and reproducibility.

### Adaptive Inference Under Limited Data

TumorCLIP supports a range of data-efficient inference modes by decoupling visual feature extraction from semantic reasoning. In the zero-shot scenario, predictions are based solely on text prototypes derived from radiology-informed prompts, with the DenseNet121 backbone generating image embeddings that are directly matched to these class-level semantic anchors. This approach allows for effective classification without any labeled training data, enabling meaningful decision-making in fully unsupervised settings.

When limited labeled data are available, TumorCLIP leverages instance-level visual evidence through a non-parametric feature cache. Training image features are stored once in this cache, and during inference, visually similar examples are retrieved to complement predictions informed by the text prototypes. This cache-guided refinement enhances decision boundaries for underrepresented classes while maintaining the frozen state of both visual and text encoders.

For cases requiring greater adaptability, a lightweight trainable adapter can be introduced to better align the visual embedding space with both cache-derived features and text prototypes. By restricting trainable parameters to this adapter, the multimodal design remains computationally efficient.

Collectively, these inference strategies define a unified continuum of adaptation—from zero-shot semantic reasoning, to training-free instance-based retrieval, to targeted lightweight tuning—offering a flexible framework that can in principle accommodate diverse data availability scenarios.

### Evaluation Metrics

Model performance was evaluated using established multi-class assessment metrics. To address class imbalance among the six diagnostic categories, we primarily report overall accuracy. In addition, class-wise performance was analyzed using confusion matrices to characterize sensitivity patterns and subtype-level misclassification across tumor categories. For external dataset evaluation, all metrics were computed using the same definitions, and performance was assessed under a filtered protocol restricted to tumor categories shared across datasets. Both zero-shot and few-shot evaluation protocols mirrored those of the fully fused TumorCLIP model, with the sole distinction being whether cache-derived logits were included or excluded during inference. No task-specific thresholds or post-processing adjustments were applied, ensuring comparability across evaluation modes. To qualitatively assess class separability in the decision space, t-SNE was applied to the learned feature embeddings extracted from the final layer. This visualization served as a complementary tool to evaluate cluster compactness and the distinctness of inter-class boundaries. Unless otherwise noted, reported results correspond to the model checkpoint achieving the highest validation accuracy across all training epochs, and test metrics are derived from this selected checkpoint.

All hyperparameters are fixed across experiments. Evaluation on the external dataset is conducted without any fine-tuning or adaptation.

### Software

All models were implemented using PyTorch. The CLIP text encoder was initialized with publicly available pretrained weights. Image preprocessing employed standard Python libraries, and no custom CUDA kernels or proprietary implementations were utilized.

## Discussion

This work presents TumorCLIP, a lightweight vision-language framework developed to address persistent limitations in deep learning-based brain tumor MRI classification. Through a systematic benchmark of eight visual backbones under a unified hyperparameter grid, DenseNet121 was identified as a robust and stable foundation for multimodal extension. Building on this backbone, TumorCLIP incorporates frozen clinical text descriptions via a Tip-Adapter mechanism, conferring several advantages.

Firstly, TumorCLIP advances explainability through concept-level reasoning, as text prototypes derived from radiological descriptions anchor model predictions in human-interpretable semantics, which is a functionality lacking in vision-only models. This is empirically supported by the semantic margin analysis, in which TumorCLIP achieves a positive margin rate of 96.9% versus 42.1% for the DenseNet baseline, confirming that alignment is not an implicit byproduct of accurate classification but requires explicit vision-language integration. Secondly, the complete framework achieves competitive classification performance, while the text-prototype space provides a complementary representation-level view of class semantics. Thirdly, TumorCLIP is computationally efficient: by freezing the CLIP text encoder and training only a lightweight adapter, training costs are lower than those of fully end-to-end multimodal architectures. Lastly, the framework provides intrinsic zero-shot and few-shot capabilities, highlighting its deployment flexibility in scenarios with limited labeled data.

Beyond in-domain evaluation, additional validation on an independent external dataset demonstrates TumorCLIP’s robustness to cross-dataset variability. While both DenseNet121 and TumorCLIP degrade in performance when evaluated on data from a different source, the decline observed for DenseNet121 is more pronounced. This effect is especially evident for glioma, a highly heterogeneous and infiltrative tumor type whose imaging appearance is sensitive to variations in acquisition protocols and scanner characteristics. Conventional CNNs may therefore over-rely on dataset-specific visual cues, leading to degraded performance when such cues shift across datasets. In contrast, TumorCLIP aligns visual representations with radiology-informed semantic concepts, encouraging reliance on diagnostically relevant pathological patterns rather than superficial imaging style. This multimodal semantic constraint contributes to the improved robustness and generalization observed across datasets. Prototype similarity captures class-level semantic correspondence but should not be interpreted as evidence of case-specific explanation fidelity.

Our findings underscore that integrating structured radiology knowledge can improve the reliability and clinical interpretability of MRI-based tumor classification systems in our experiments. Future research will seek to broaden the diversity of text prototypes, incorporate structured radiology ontologies, and validate the model’s generalizability across multi-institutional datasets. A limitation of this study is the reliance on manually authored radiology-style text prototypes, which may introduce variability depending on expert knowledge and reporting conventions. In addition, differences in imaging protocols or institutional practices may affect the transferability of learned semantic alignments across datasets.

## Author Contributions

Y.J. and J.N. conceived the study, trained the models, performed data analysis, and prepared the manuscript and figures. J.G. supervised the project, designed the study, defined the scope of the work, and contributed to the overall content of the manuscript. A.L. reviewed and supervised the manuscript. Z.Q. and Z.L. revised the manuscript. All authors approved the final manuscript.

## Code Availability

The code used in this study is publicly available at:

https://github.com/3o30/TumorCLIP

## Data Availability

The dataset used in this study is publicly available at: https://www.kaggle.com/datasets/fernando2rad/brain-tumor-mri-images-17-classes, https://www.kaggle.com/datasets/waseemnagahhenes/brain-tumor-for-14-classes

## Data Availability

The datasets used in this study are publicly available.

https://www.kaggle.com/datasets/fernando2rad/brain-tumor-mri-images-17-classes

https://www.kaggle.com/datasets/waseemnagahhenes/brain-tumor-for-14-classes

