## Supplementary Table 1 for "TumorCLIP: Radiology-informed vision-language alignment for interpretable MRI-based brain tumor classification"

### S1: Tumor type merge table

Fernando2rad. Brain Tumor MRI Images (17 Classes)

| Main category | Sub Category |
|---|---|
| Glioma | Astrocytoma, Ganglioglioma, Glioblastoma, Oligodendroglioma, Ependymoma |
| Meningioma | Low Grade, Atypical, Anaplastic, Transitional |
| Neurocytoma | Central - Intraventricular, Extraventricular |
| NORMAL | |
| Outros Tipos de Lesões | Abscesses, Cysts, Miscellaneous Encephalopathies |
| Schwannoma | Acoustic, Vestibular - Trigeminal |

Nagahhenes, W. Brain Tumor for 14 Classes

| Main category | Sub Category |
|---|---|
| Glioma | Astrocytoma, Ganglioglioma, Glioblastoma, Ependymoma |
| Meningioma | Meningioma |
| Neurocytoma | Neurocytoma |
| NORMAL | |
| Schwannoma | Schwannoma |
